# “Distribution, genetic polymorphism and genotype prediction of rhesus blood group antigens among the Kurdish population of Zakho, Kurdistan Region, Iraq”

**DOI:** 10.1101/2025.11.20.25340675

**Authors:** Shakir A. Zebari, Sawer Ahmed, Ibrahim Naqid, Shivan Muhammed, Abdullah Muhi

## Abstract

The Rhesus blood group system exhibits significant polymorphism, with diverse antigen distributions across populations. This study investigates the antigenic profile, haplotype frequencies, and genotype predictions in a cohort from Zakho, Kurdistan Region, Iraq, which is crucial for transfusion medicine and genetic studies. A prospective cross-sectional analysis was performed on 1,000 Kurdish individuals at Zakho Emergency Teaching Hospital. Blood samples were phenotyped for Rh (D, C, c, E, e) antigens. Haplotype assignments were made using Fisher-Race terminology, and probable genotypes were calculated based on allele frequencies and Wiener nomenclature. The most common Rhesus blood antigens are Rh(e) (95%), Rh(D) (91.9%), Rh(C) (76%), and Rh(c) (68%). Rh(E) is found in 25.8% of individuals, while only 8.1% are Rh(D) negative. Among the Rh(D) positive population, the most frequent phenotype/haplotype and presumed genotype was DCe (*DCe/DCe, R1R1*) at 31.4%, followed by DCce (*DCe/dce, R1r*) at 29.3%, and DCcEe (*DCe/DcE, R1R1*) at 13.8%. The dce phenotype (*dce/dce, rr*, in 7.2%) was the most common among Rh(D) negative individuals. No significant differences were observed between sexes. This study reveals that DCe, DCce, and DCcEe are prevalent phenotypes among Rh(D) positive individuals, whereas the dce haplotype predominates among Rh(D) negative individuals. The Rhesus phenotype/genotype aligns with Kurdish and Arab groups in Iraq and shows partial resemblance to the Western European descent, Indian, and Iranian populations, but significantly differs from African-American populations, except for the dce phenotype (*dce/dce, rr)*. These findings are important for blood transfusion strategies, donor selection, and genetic research in the region.

## Introduction

The Rhesus blood group system is the second most frequently considered blood group system in clinical practice, following the ABO system. [1]. The Rh blood group is encoded by two tightly linked loci located on chromosome 1; specifically, the RHD gene encodes the RhD antigen, while the RHCE gene encodes the RhCE antigens. A sequence of unknown significance known as *SMP1* separates the *RHD* and *RHCE* genes [2]. The Rh blood group system exhibits considerable complexity, as various antigens arise from combinations of single-nucleotide variations and gene rearrangements [3].

This blood group system comprises 55 antigens, which are carried on two proteins, RhD and RhCE, each consisting of 417 amino acids. However, only five antigens (D, C, c, E, and e) are recognised as clinically significant. Among these, the Rh (D) antigen is noteworthy due to its highly immunogenic properties, leading to extensive research and testing. This is largely attributed to its association with hemolytic transfusion reactions [4]. The lack of compatibility testing for Rh blood groups may result in the development of antibodies during transfusion; the recipient’s immune system produces such antibodies in response to foreign Rh antigens present on transfused red blood cells (RBCs). These immune responses can precipitate serious clinical complications, including acute hemolysis, organ dysfunction, or fetal morbidity in Rh-negative individuals [5].

Numerous studies in Iraq have documented the distribution of ABO and Rh(D) blood groups among Kurdish and Arab populations. [6,7]. Data on the frequencies of other Rh antigens (C, c, E, e) are scarce. The present study was conducted to evaluate the prevalence, phenotypic distribution, and genotypic prediction of the Rhesus blood group system within the Kurdish population residing in Zakho City, Kurdistan Region of Iraq.

## Material and methods

This prospective cross-sectional study was conducted at a premarital screening centre at Zakho Emergency Teaching Hospital between January 2021 and January 2023. The study included the first five couples who attended mandatory premarital screening daily, with a total of 1000 participants included in this study.

Non-Kurdish populations and non-residents of Zakho City were excluded from this study. Verbal informed consent was obtained from all participants. Sociodemographic data were collected, including names, dates of birth, residency, and phone numbers. These records were stored and kept in confidential files and accessed only by authorised personnel.

Three millilitres (ml) of blood were collected from each participant by proper phlebotomy techniques. The blood was distributed into a K3-Ethylene-Diamine-Tetraacetic-Acid (EDTA) vacutainer tube and well mixed on a rotatory mixer.

The Rh(D) antigen status of all participants was determined using the standard slide agglutination technique with BIOSCOT CE-marked IgM monoclonal anti-D reagent (Bioscot Ltd., United Kingdom, distributed in the U.S. and Canada). Additionally, phenotyping for the other Rh antigens (C, c, E, and e) was performed using the conventional tube agglutination method with the corresponding BIOSCOT^®^ CE-marked IgM monoclonal antisera (anti-C, anti-c, anti-E, and anti-e), from Bioscot Ltd. All procedures were conducted in strict accordance with the manufacturer’s instructions.

Phenotype frequencies of Rhesus antigens were calculated by dividing the count of positive each antigen by the total number of individuals screened, with results expressed as a percentage. Five main Rhesus antigens were evaluated using antisera D, C, E, c, and e; phenotypes and genotypes were designated according to Fisher-race/ Wiener’s nomenclature. Precise genotype determination is unfeasible without testing family members or conducting DNA analysis; thus, the most probable genotype is inferred from gene frequency estimates.

Descriptive statistical methods were employed utilising IBM SPSS Statistics, version 27, to elucidate the findings of the study, including a Chi-square test of independence, with a p-value threshold of less than 0.05 deemed statistically significant.

## Results

From all 1000 enrolled subjects, 500 (50%) were males and 500 (50%) were females. The median age group was 25.4 years, ranging from 18 to 55 years.

The overall prevalence of Rh(D) positive was found in 919 (91.9%) of subjects, while 81 (8.1%) were found to be Rh(D) negative. No statistically significant difference in Rh(D) distribution was observed among males and females, “Table 1”.

**Table 1.** Distribution of Rh(D) antigen status by sex among the studied population.

| Sex | Rh(D) Positive n (%) | Rh(D) Negative n (%) | Total n (%) | *p-value |
| --- | --- | --- | --- | --- |
| Female | 460 (92.0) | 40 (8.0) | 500 (50) | 1.0 |
| Male | 459 (91.8) | 41 (8.2) | 500 (50) |  |
| Total | 919 (91.9) | 81 (8.1) | 1000 (100) |  |

Variable distribution of other Rhesus blood antigens was observed among the studied population. Rh(e) antigen was present in 960 (96%), followed by Rh (C) antigen in 761 (76.1%) and Rh(c) antigen in 680 (68%), while Rh (E) antigens were the least prevalent and were only positive in 258 (25.8%). No statistically significant difference in Rhesus (e, C, c, E) antigen distribution was observed among males and females, “Table 2”.

**Table 2.** Distribution of other Rhesus blood group antigens by sex among the studied population.

| Rhesus Phenotype | Overall Positive n (%) | Male n (%) | Female n (%) | * *p* -value |
| --- | --- | --- | --- | --- |
| **Rh(e)** | 960 (96.0%) | 481 (96.2%) | 479 (95.8%) | 0.82 |
| **Rh(C)** | 761 (76.1%) | 381 (76.2%) | 380 (76.0%) | 0.94 |
| **Rh(c)** | 680 (68.0%) | 340 (68.0%) | 340 (68.0%) | 1 |
| **Rh(E)** | 258 (25.8%) | 128 (25.6%) | 130 (26.0%) | 0.89 |

In this study, the most prevalent Rhesus phenotypes/ Haplotypes were DCe (31.4%), DCce (29.3%), and DCcEe (13.8%). These were followed by DcEe (8.0%), dce (7.2%), Dce (5.4%), and DcE (3.3%). Less frequent phenotypes/Haplotypes included DCcE (0.7%), dCe (0.6%), and Cce (0.3%). A variety of phenotypes, notably (DCEe, DCE, CcEe, CcE, CE, cEe and cE) were not detected in this cohort “Table 3”.

**Table 3.** Frequency of Rhesus blood group antigens and corresponding Fisher-Race haplotypes/phenotype distribution among the studied population.

| *Reaction pattern with antisera |  |  |  |  | Fisher–Race<br>haplotype /<br>Phenotype | Number of<br>Cases (n) | Percentage<br>(%) |
| --- | --- | --- | --- | --- | --- | --- | --- |
| D | C | c | E | e |  |  |  |
| + | + | – | – | + | DCe | 314 | 31.4 |
| + | + | + | – | + | DCce | 293 | 29.3 |
| + | + | + | + | + | DCcEe | 138 | 13.8 |
| + | – | + | + | + | DcEe | 80 | 8 |
| + | – | + | – | + | Dce | 54 | 5.4 |
| + | – | + | + | – | DcE | 33 | 3.3 |
| + | + | + | + | – | DCcE | 7 | 0.7 |
| – | – | + | – | + | ce | 72 | 7.2 |
| – | + | – | – | + | Ce | 6 | 0.6 |
| – | + | + | – | + | Cce | 3 | 0.3 |
| Total |  |  |  |  |  | 1000 | 100 |

\*(+) indicate positive reaction, (-) indicate negative reaction.

When these results are stratified by Rh(D) status, it is observed that among Rh(D) positive samples, the most common phenotypes/haplotype are DCe (34.2%), DCce (31.9%), DCcEe (15%), followed by DcEe (8.7%), Dce (5.9%), DcE (3.6%), and DCcE (0.8%). Reflecting the predominant haplotype distribution in Rh(D+) individuals. While among Rh(D) negative samples, the dce haplotype was the most predominant (88.9%), followed by dCe (7.4%) and Cce (3.7%), highlighting the dominance of the dce haplotype among Rh(D–) individuals.

In this study, the most probable genotypes among Rh(D) positive individuals were *DCe/DCe (R*_*1*_*R*_*1*_, 34.2%), *DCe/dce (R*_*1*_*r*, 31.9%), and *DCe/DcE (R*_*1*_*R*_*2*_, 15.0%). Less frequent genotypes included *DcE/dce (R2r*, 8.7%), *Dce/dce (R0r*, 5.9%), *DcE/DcE (R*_*2*_*R*_*2*_, 3.6%), and *DCE/DcE (RzR*_*2*_, 0.8%) “Table 4”. Among Rh(D) negative individuals, the predominant genotype was *dce/dce (rr*, 88.9%), followed by *dCe/dce (r’r*, 7.4%) and *dCe/dCe (r’r’*, 3.7%) “Table 4”.

**Table 4.**
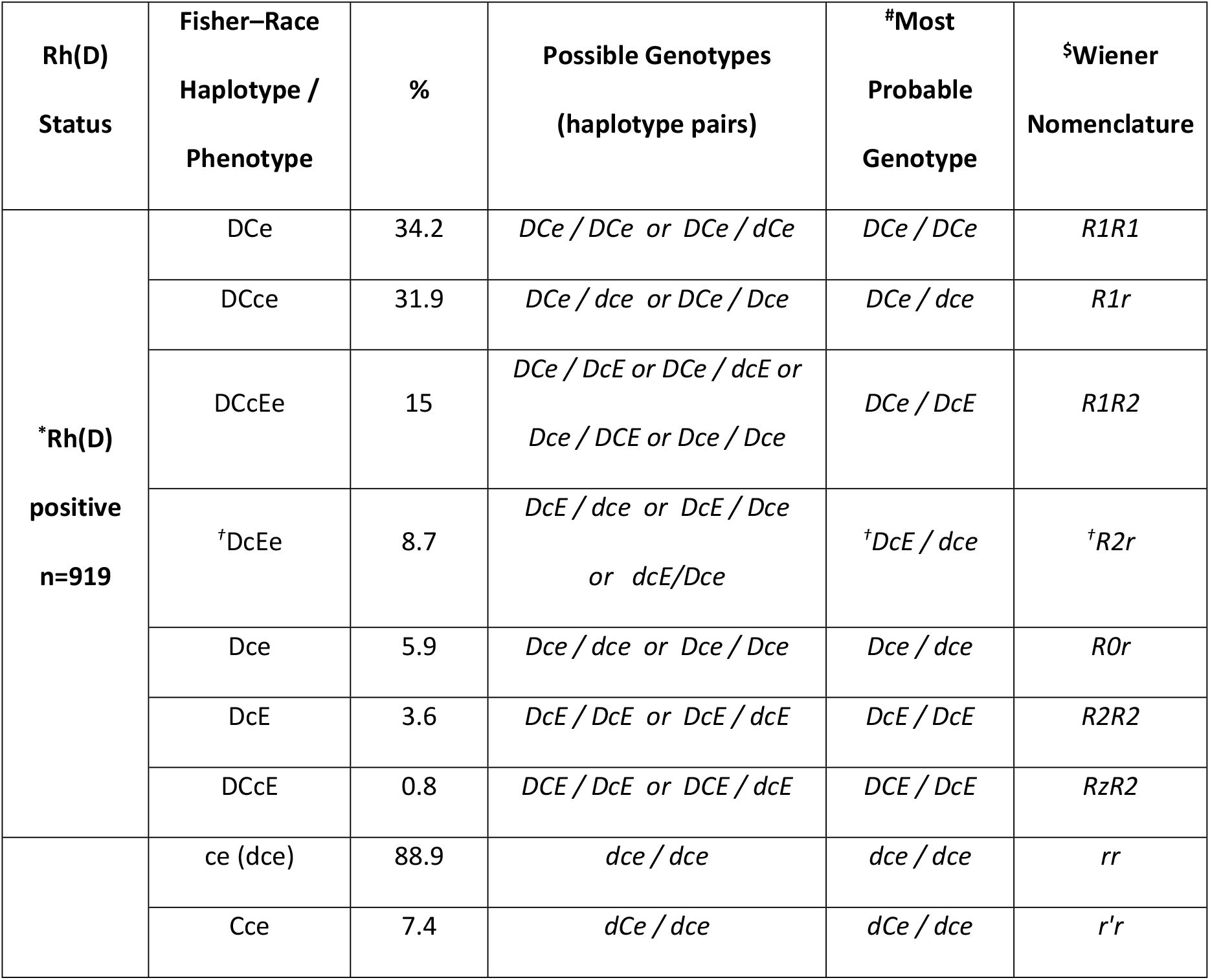
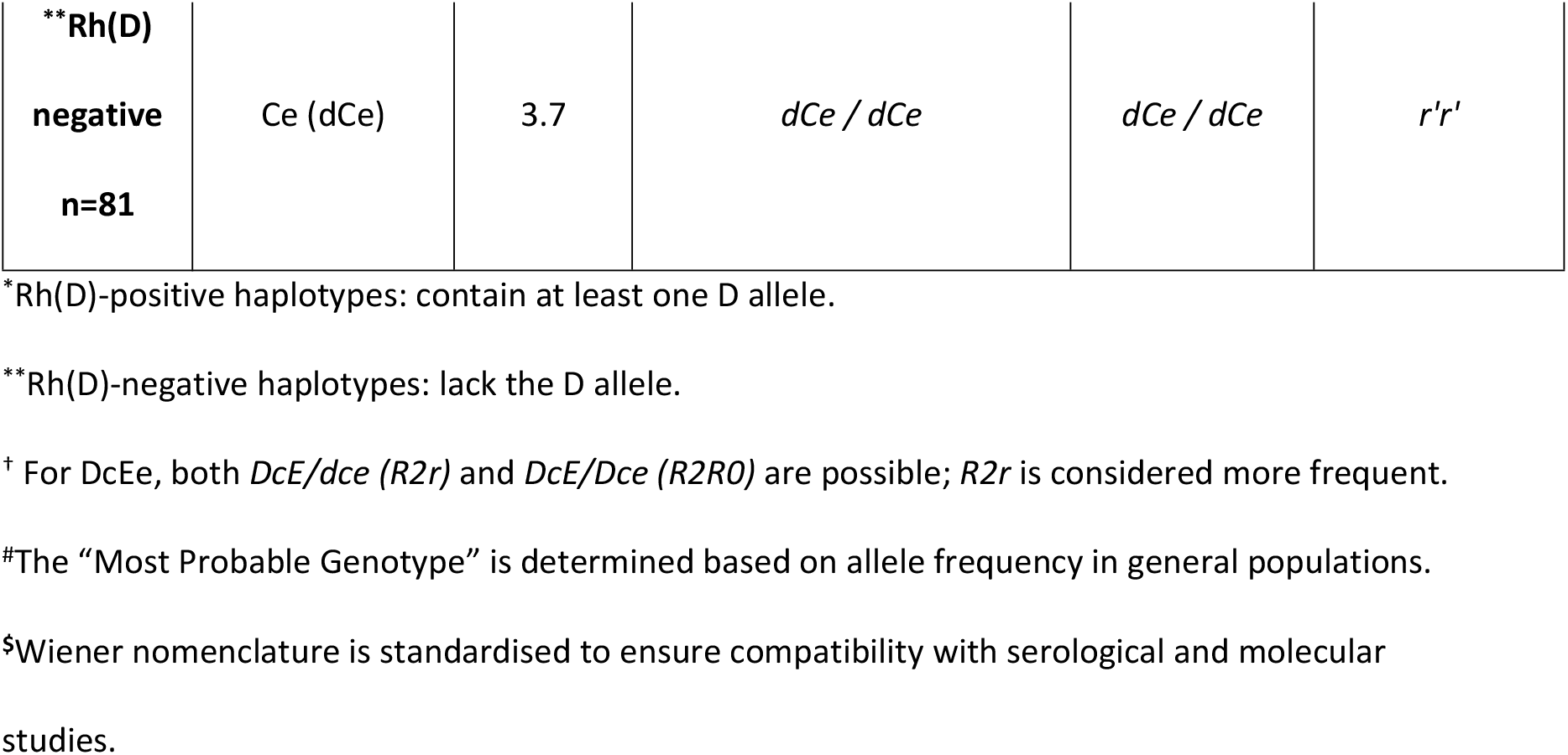
Fisher-Race haplotype/Phenotype distribution, Genotype Prediction, most probable genotype and Wiener Nomenclature by Rh(D) Status among the studied population.

## Discussion

The study of blood group systems provides insights into human evolution and diversity. Analysing variations among populations helps trace migrations, reveal genetic relationships, and understand adaptations over time. Additionally, knowledge of blood group distribution is crucial in clinical medicine, genetic research, and anthropology, linking biology to our shared heritage [8].

Blood transfusions are essential for saving lives but carry risks. The immune system may produce alloantibodies when exposed to incompatible red blood cells, leading to reactions that can range from mild to severe, including hemolytic disease in fetuses and hemolytic reactions post-transfusion [9].

Antibodies against clinically significant antigens, like Rhesus antigens, can cause hemolytic transfusion reactions and hemolytic disease in newborns. These complications may be encountered routinely in clinical practice. Recognising these complications is essential in clinical practice, and comprehensive phenotypic detection of these antigens can help in their prevention and management [7].

The Rh antigen is inherited in a codominant manner via the autosomal chromosome 1, suggesting no significant sex differences. However, due to the Kurdish population’s diverse religious groups (Muslim, Christian, and Yezidi), comparative analyses were performed to account for variations in Rh antigen polymorphisms linked to religious differences.

The current study revealed that the distribution of the Rh(D)positive antigen was 91.9%. This finding aligns with that from a previous study in Iraq-Duhok [6], suggesting genetic homogeneity among Kurdish populations in northern Iraq, and with that of the African-American [13]. However, the observed prevalence is lower than that recorded in China [11], and it is slightly higher than the prevalence found within the Iraqi-Arab, Iranian, Indian and Western European descent population [7.10.15.13] “Table 5”.This may be due to geographical variation and ethnic background of the population.

The Rh(D) negative frequency in this study (8.9%) aligns closely with Iraqi-Duhok [6] and African-American [13], but is lower than reported from other global studies [7,10-15]. The similarity between Zakho and Duhok suggests genetic homogeneity among Kurdish populations in northern Iraq. In contrast, Basrah (southern Iraq, more Arab-majority) shows a slightly higher Rh(D) frequency, possibly reflecting historical gene flow from neighbouring Persian or South Asian populations. The low Rh(D)-negative prevalence rate underscores the need for a D-negative blood inventory for emergency, obstetric, and transfusion services.

The frequency of the Rh(e) antigen observed in this study is consistent with findings reported in various global studies [6,7,10-15]. The e allele is recognised as the ancestral and most prevalent variant at the RHCE locus. Typically, frequencies of the Rh(e) antigen exceed 95% worldwide. Consequently, the significant prevalence of the Rh(e) antigen poses considerable challenges in identifying e-negative blood donors for patients with alloanti-e antibodies. Furthermore, the occurrence of anti-e as an autoantibody adds another layer of complexity to the provision of compatible blood, “Table 5”.

**Table 5.** Comparative frequencies (%) of Rhesus antigens among different populations.

| Population / Region | Rh(D)+ | Rh(D)– | Rh(C) | Rh(c) | Rh(E) | Rh(e) |
| --- | --- | --- | --- | --- | --- | --- |
| Iraq (Duhok) [6] | 91.1 | 8.9 | 75.9 | 68.1 | 25.1 | 95.6 |
| Iraq (Basrah) [7] | 88.8 | 11.2 | 68.3 | 77.7 | 52.6 | 96.7 |
| Iran [14] | 87.76 | 12.24 | 73.6 | 72.1 | 30.83 | 93.59 |
| China [11] | 99.4 | 0.6 | 88.77 | 53.63 | 44.94 | 92.61 |
| India [15] | 84.35 | 15.65 | 81.74 | 56.32 | 21.74 | 100 |
| Western European descent [13] | 85 | 15 | 70 | 80 | 30 | 98 |
| African-American [13] | 92 | 8 | 34 | 97 | 21 | 99 |
| Current study | 91.1 | 8.9 | 76.1 | 68 | 25.8 | 96 |

The prevalence of the Rh(E) antigen in this study is consistent with findings reported among Kurdish populations in Duhok, Iraq [6], and in certain regions of Iran [10], suggesting a limited frequency of E-carrying Rh haplotypes (e.g., DcE, dcE). However, it is higher than the frequencies reported in Indian and African-American populations [12,13], and lower than those observed in Arab populations in Basra, Iraq [7], as well as in Chinese and Western European descent populations [11,13]. Despite its relatively low population prevalence, the Rh(E) antigen is highly immunogenic, second only to D among Rh antigens. Consequently, anti-E is among the most frequently encountered non-D Rh antibodies in multiply transfused patients. This may explain the higher rates of alloimmunization to E compared with the more prevalent but less immunogenic Rh(e) antigen “Table 5”.

In this study, the high frequency of Rh(C) antigen mirrors that of Kurdish populations in Duhok, Iraq [6], Iran [10], and India [12], suggesting shared Indo-Iranian or ancient Mesopotamian genetic influences. Furthermore, there is some degree of resemblance to the Iraqi-Arab and Western European descent population [7,13]. However, the levels observed in our study are lower than those documented for the Chinese [11] and significantly higher than those found in the African-American population [13] “Table 5”.

Regarding the Rh(c) antigen, our study shows similarity to that reported from Kurdish Iraqis [6] and to some extent with the Iranians [10] and slightly lower than the Iraqi Arabs (Basrah) [7], possibly indicating regional substructure within Iraq. While it is quite higher than reported by the Chinese and Indian [11,12], and lower than reported from Western European descent and African-American [13], “Table 5”.

This study evaluated Rh phenotype frequencies based on observed antigen expression patterns and established genotype–phenotype correlations from previous population studies. Direct genotyping was not conducted; probable genotype assignments were made following documented allelic associations from earlier population studies [13].

The most frequent Rhesus haplotype observed among the Rh(D)-positive Kurdish population in this study was DCe (R_1_), followed by Dce (R_0_) and DcE (R_2_), corresponding to the presumed genotypes *R*_*1*_*R*_*1*_, *R*_*1*_*r*, and *R*_*1*_*R*_*2*_, with frequencies of 31.4%, 29.3%, and 13.8%, respectively. Among the Rh(D)-negative individuals, the most prevalent haplotype was *ce* (*r*), corresponding to the *rr* genotype, which accounted for 7.2% of the total studied population “Table 6”.

**Table 6.** Comparative frequencies of common Rhesus genotypes in different populations.

| Rh(D)<br>Status | Genotype<br>(Fisher-Race) | Iraq<br>(Duhok)[6] | Western<br>European<br>descent [13] | African-American<br>[13] | Indian<br>[15] | Iranian<br>[14] | Current<br>Study |
| --- | --- | --- | --- | --- | --- | --- | --- |
| Positive | *R1R1 (DCe/DCe)* | 31.5 | 18.5 | 3 | 40.87 | 28.23 | 31.4 |
|  | *R1r (DCe/Dce)* | 30.1 | 34.9 | 21 | 23.48 | 26.22 | 29.3 |
|  | *R1R2 (DCe/DcE)* | 13.8 | 13.3 | 4 | 13.91 | 12.74 | 13.8 |
| Negative | *rr (dce/dce)* | 7.4 | 15.1 | 7 | 11.3 | 9.02 | 7.2 |

Data are presented as percentages.

The *R1R1* genotype is notably high in the current study, showing close similarity to findings previously reported from Duhok, Iraq [6], and, to some extent, with the Iranian population [14], but is much higher than the Western European and African-American frequencies [13]. It is slightly lower than in Indian populations [15]. This suggests a shared genetic background within the region, possibly reflecting common ancestry or limited admixture with populations where R1R1 is less common (e.g., African or European groups) “Table 6”.

The *R1r* frequency in the current study is similar to that reported from the Duhok Iraqi population [6], indicating internal consistency. It is lower than in the Western European descendants [13] but higher than in African-American, Iranian and Indian groups [13-15]. Suggesting this genotype is stable, may be ancestral or selectively neutral across a wide geographic range, but less common in those with predominant African ancestry, “Table 6”.

The *R1R2* genotype is notably high in the Kurdish population, closely mirroring the broader Iraqi Duhok, the Western European descent and Indian [6,13,15]; however, it is much higher than reported from African-American [13] and lower than the Iranian populations [14] “Table 6”.

The *dce/dce (rr*, 7.2%) genotype frequency reported in this study is consistent with global patterns of the Rh(D) negative population; however, it is among the lowest reported, similar to African-Americans and the Iraqi Kurdish population [6, 13] and slightly lower than reported from Iran and India [14,15]. This contrasts sharply with the Western European descents, where Rh negativity is nearly twice as common [13]. The similarity with African-Americans may be coincidental, as the underlying haplotype structures differ. The lower Rh(D)-negative prevalence in Kurds implies reduced risk of hemolytic disease of the fetus and newborn (HDFN) compared to the Western European descent populations, but antenatal screening remains essential, “Table 6”.

## Conclusion

The findings indicate that the prevalence of Rh system antigens and phenotypes varies among different geographical regions, highlighting the need for regional studies. While blood transfusion centres routinely test for ABO and Rh(D) blood types, the additional Rh antigens (C, c, E, e) are not consistently screened, posing a risk of alloimmunization for recipients lacking these antigens. This study serves as a preliminary effort to identify the dominant Rh blood group system among the Kurdish population in Zakho city to improve healthcare services and better meet patient needs.

The current study confirms that e, D, C, and c antigens are common in the Rhesus blood group system, with the E antigen present in 25.8% of the population and only 8.1% of subjects were identified as Rh(D) negative.

This study reveals that DCe, DCce, and DCcEe are prevalent phenotypes among Rh(D) positive individuals, whereas the ce haplotype predominates among Rh(D) negative individuals. The Rhesus phenotype/genotype aligns with Kurdish and Arab groups in Iraq and shows partial resemblance to the Western European descent, Indian, and Iranian populations, but significantly differs from African-American populations, except for the dce phenotype (*dce/dce, rr)*.

Our understanding of the Rh blood group system now extends to transfusion-related issues, alloimmunization in multi-transfused patients, and disease associations with red blood cell surface antigens. Increased awareness of blood group diversity will improve blood bank management and volunteer donor registries. This research is vital and should be replicated in other regions of Kurdistan and Iraq to address challenges related to hemolytic transfusion reactions and hemolytic disease of the fetus and newborn.

## Data Availability

All relevant data are within the manuscript

## Acknowledgement

The authors would like to acknowledge the staff of the premarital screening centre at Zakho emergency teaching hospital for their cooperation.

## Notes

### Competing Interest Statement

The authors have declared no competing interest.

### Funding Statement

The author(s) received no specific funding for this work.

### Author Declarations

The study was reviewed and approved by the Research Ethics Committee, College of medicine, University of Zakho, Zakho, Kurdistan Region, Iraq

